# Kinetics of the humoral immune response to SARS-CoV-2: comparative analytical performance of seven commercial serology tests

**DOI:** 10.1101/2020.06.09.20124719

**Authors:** Pauline H. Herroelen, Geert A. Martens, Dieter De Smet, Koen Swaerts, An-Sofie Decavele

## Abstract

**Background:** SARS-CoV-2 serology tests are clinically useful to document a prior SARS-CoV-2 infection in patients with no or inconclusive PCR results and suspected COVID-19 disease or sequelae. Data are urgently needed to select the assays with optimal sensitivity at acceptable specificity.

**Methods:** A comparative analysis of analytical sensitivity was performed of seven commercial SARS-CoV-2 serology assays on 171 sera from 135 subjects with PCR-confirmed SARS-CoV-2 infection, composed of 71 patients hospitalized for COVID-19 pneumonia and 64 healthcare workers with paucisymptomatic infections. The kinetics of IgA/IgM/IgG seroconversion to viral N-and S-protein epitopes were studied from 0 to 54 days after symptom onset. Specificity was verified on 57 pre-pandemic samples.

**Results:** Wantai SARS-COV-2 Ab ELISA and Orient Gene COVID-19 IgG/IgM Rapid Test achieved a superior overall sensitivity. Elecsys Anti-SARS-CoV-2 assay and EUROIMMUN Anti-SARS-CoV-2 combined IgG/IgA also showed acceptable sensitivity (>95%) versus the consensus result of all assays from 10 days post symptom onset. Optimal specificity (>98%) was achieved only by Wantai SARS-COV-2 Ab ELISA, Elecsys Anti-SARS-CoV-2 assay and Innovita 2019-nCoV Ab rapid test. LIAISON SARS-CoV-2 S1/S2 IgG showed a significantly lower sensitivity as compared to all other assays. Lack of seroconversion by any test was seen in 1.4% of hospitalized and 4.7% of paucisymptomatic infections. Within 10 days from symptom onset, only the Wantai SARS-COV-2 Ab ELISA has acceptable sensitivity.

**Conclusions:** Wantai SARS-COV-2 Ab ELISA and Elecsys Anti-SARS-CoV-2 assays are suitable for sensitive and specific screening of a SARS-CoV-2 infection from 10 days after symptom onset.

**Brief summary:** There is an urgent need for SARS-CoV-2 serology tests for the sensitive and specific detection of prior SARS-CoV-2 infection as a complementary diagnostic tool to molecular testing. Various commercial assays are becoming available but comparison of their relative performance is difficult unless they are head-to-head evaluated. Here we compared seven commercial assays on sera equally composed of mild and severe PCR-confirmed SARS-CoV-2 infections. Our analysis indicates a superior performance of the Wantai SARS-COV-2 ELISA for total antibodies to the S-RBD domain. Also, the Elecsys Anti-SARS-CoV-2 assay for total antibodies to the N-protein shows good performance for high-throughput screening.

## Introduction

The gold standard for diagnosis of COVID-19 lung disease is nucleic acid amplification testing of SARS-CoV-2 virus-specific sequences coding for the spike (S), envelope (E) and nucleocapsid (N) proteins, the RNA dependent RNA polymerase (RdRp) gene and the Open Reading Frame 1ab (ORF1ab) region (1). The diagnostic sensitivity of the most commonly used technique, RT-PCR on nasopharyngeal swabs is currently unknown. When compared to chest CT analysis of lesions characteristic for viral pneumonia, estimates vary from lower than 70% to 90% (2,3) likely depending on COVID-19 disease stage, the intensity of viral replication, sampling quality and analytical properties of the amplification assay. In addition, insufficient PCR capacity during peak infection rate in overwhelmed healthcare systems left many patients with milder clinically suspected SARS-CoV-2 infections as well as asymptomatic infections untested.

Serology testing for COVID-19, defined as the detection of IgM, IgA or IgG antibodies to SARS-CoV-2-specific epitopes, might represent an interesting complementary diagnostic tool to document a past SARS-CoV-2 infection, both in individual patients with suspected COVID-19 symptoms or late-stage complications who had no (conclusive) PCR test as at population level to guide infection control policies. In addition, measuring SARS-CoV-2 antibodies might harbor prognostic value and convey information on protective immunity in vaccination trials.

SARS-CoV-2 is an enveloped, single-stranded RNA betacoronavirus (βCoV), belonging to the *Coronaviridae* family. All human coronaviruses share four major structural proteins: envelope, membrane, nucleocapsid, and spike protein. SARS-CoV-2 shares a 80% overall nucleotide homology with SARS-CoV (4–6). In SARS-CoV, the spike- and nucleocapsid protein contain the highest density of B-cell epitopes (7,8) and in silico analysis indicated that dominant B-cell epitopes share 69% to 100% homology to SARS-CoV-2. It was therefore a logical choice of many commercial developers of diagnostic SARS-CoV-2 serology kits to target the S and N proteins. In addition to their diagnostic value, antibodies to the S protein, composed of a S1 subunit with the receptor binding domain (RBD) and a S2 subunit that mediates membrane fusion for viral entry, appear additionally interesting because of their proposed correlation with neutralizing antibodies and protective immunity to both SARS-CoV (6) and, based on emerging data, also to SARS-CoV-2 (9,10).

Data on the kinetics of the humoral immune response to SARS-CoV-2 are rapidly emerging but questions remain as to the relative diagnostic sensitivity of various commercial assays. In this study we present a cross-platform comparison of seven commercially available SARS-CoV-2 serology assays, targeting both N and S protein epitopes and different combinations of antibody isotypes in PCR-confirmed COVID-19 patients both with critical and mild disease course at various time points. To select assays suitable for screening the general population, we used as working definition of acceptable performance a sensitivity > 95% versus the consensus result of all tests also in mild SARS-CoV-2 infections from 10 days after onset of symptoms and a minimal specificity of 98%.

## Methods

### Patients

This is a diagnostic accuracy study on serum samples obtained from the following cohorts: (i) Hospitalized COVID-19 patients: 105 serum samples obtained at different time points from 71 patients with PCR-confirmed SARS-CoV-2 infection and admitted for severe COVID-19 pneumonia from March 1 to April 27, 2020 at AZ Delta General Hospital in Roeselare, Belgium; (ii) Paucisymptomatic SARS-CoV-2 infections: 66 serum samples from 64 healthcare workers with a SARS-CoV-2 infection, PCR-confirmed after developing fever and World Health Organization (WHO)-listed COVID-19 symptoms. These patients were home-quarantined without the need for hospitalization; (iii) Suspected SARS-CoV-2 infection: 84 serum samples from 84 healthcare workers from AZ Delta General Hospital, Roeselare and Sint-Andries Hospital, Tielt who presented WHO-listed COVID-19 symptoms but were not tested by PCR mainly due to restrictive national test indications at the time. The study was approved by the AZ Delta ethical committee with a waiver of informed consent from the hospitalized COVID-19 patients (Clinical Trial Number IRB B1172020000009) and with written informed consent from participants with paucisymptomatic and suspected SARS-CoV-2 infections (Clinical Trial Number B1172020000006).

The specificity was analyzed on a panel composed of 57 pre-pandemic serum samples obtained from patients with PCR-confirmed infection by other HCoV respiratory viruses (n=7), other pathogens and viruses (n=42) or presence of auto-immune antibodies (n=8) (Supplementary data, supplementary table 1).

### SARS-CoV-2 serology assays

All serology assays were used according to the manufacturers’ protocol using the cutoffs specified in the package inserts as detailed below.

### Rapid tests

The COVID-19 IgG/IgM Rapid Test (Zhejiang Orient Gene Biotech Co., Ltd., Zhejiang, China) is a solid phase immunochromatographic assay for the qualitative detection of IgM and IgG antibodies to recombinant N- and S-proteins. The Innovita 2019-nCoV Ab Test (Innovita Biological Technology Co., Ltd., Beijing, China) is a colloidal gold lateral flow assay for the qualitative detection of IgM and IgG antibodies to undisclosed SARS-CoV-2 epitopes. Rapid tests were considered positive if a line was observed for either IgM, IgG or both. The intensity of the color was not evaluated.

### Enzyme linked immunosorbent assays (ELISA)

The Wantai SARS-COV-2 Ab ELISA (Beijing Wantai Biological Pharmacy Enterprise, Beijing, China) is a double-antigen sandwich immunoassay for the qualitative detection of all antibody isotypes (IgM, IgA, IgG) against the RBD domain of the S1 protein. Samples with a cutoff ratio (OD/CO with cutoff = mean of three blanks + 0.16) higher than 0.9 were considered positive, classifying gray zone results 0.9-1.1 as positive. Three indirect ELISAs from EUROIMMUN AG (a PerkinElmer Company, Luebeck, Germany) were tested: the Anti-SARS-CoV-2 IgG and IgA assays for semiquantitative detection of IgA and IgG antibodies against the S1 protein and Anti-SARS-CoV-2-NCP(IgG) assay for semiquantitative detection of IgG to the N protein. (cutoff = 0.8 units, classifying gray zone results 0.8-1.1 units as positive). All ELISAs were tested using the PhD™ system (Version EIA 0_16, Bio-Rad Laboratories, Inc., Hercules, California).

### Electrochemiluminescence immunoassays (CLIA)

The Elecsys Anti-SARS-CoV-2 assay for Cobas e601 module (Roche Diagnostics, Basel, Switzerland) is a double-antigen sandwich assay for the qualitative detection of all antibody isotypes (IgM, IgA, IgG) against the N protein (cutoff = 1 Cutoff Index) LIAISON SARS-CoV-2 S1/S2 IgG (DiaSorin, Saluggia, Italy) is an indirect CLIA for the quantitative detection of IgG antibodies against S1/S2 proteins (cutoff = 12 AU/mL, classifying gray zone results between 12 and 15 AU/mL as positive).

*SARS-CoV-2 PCR* was done using the Allplex™ 2019-nCoV assay (Seegene, Seoul, Korea) for E/N/RdRP genes on nasopharyngeal swab.

### Statistical analysis

Statistical analyses were performed using MedCalc (version 12.2.1, Belgium). The sensitivity of serology tests was evaluated on samples obtained from SARS-CoV-2 PCR-positive patients as (i) total fraction of samples showing detectable antibodies and (ii) by comparing each individual assay versus the consensus outcome obtained by the majority of all assays evaluated in this study. Diagnostic test (2×2) was used for calculation of sensitivity and specificity. Chi-squared (χ2) test was used for comparing proportions for categorical variables. Not-normally quantitative variables are expressed as medians (IQR) and Mann-Whitney test was used to test statistical differences between various timeframes after symptom onset. Differences were considered statistically significant if P-value was <0.05. Kinetics of seroconversion in individual patients in Fig. 1 were fitted to a scale from -1 to +1 with 0 representing each assays cutoff by subtracting each assay’s cutoff from its raw data signals, and dividing its absolute value by the highest (lowest) cutoff-corrected signal for that assay obtained in our data set for positive (negative) samples. Figure 2 was created in Python 3.7.7. The packages numpy (1.18.4), pandas (1.0.3) and matplotlib (3.2.1) were used to process the data and generate the plot. The labels were optimized using LaTeX (3.14159265-2.6-1.40.20).

**Figure 1.**
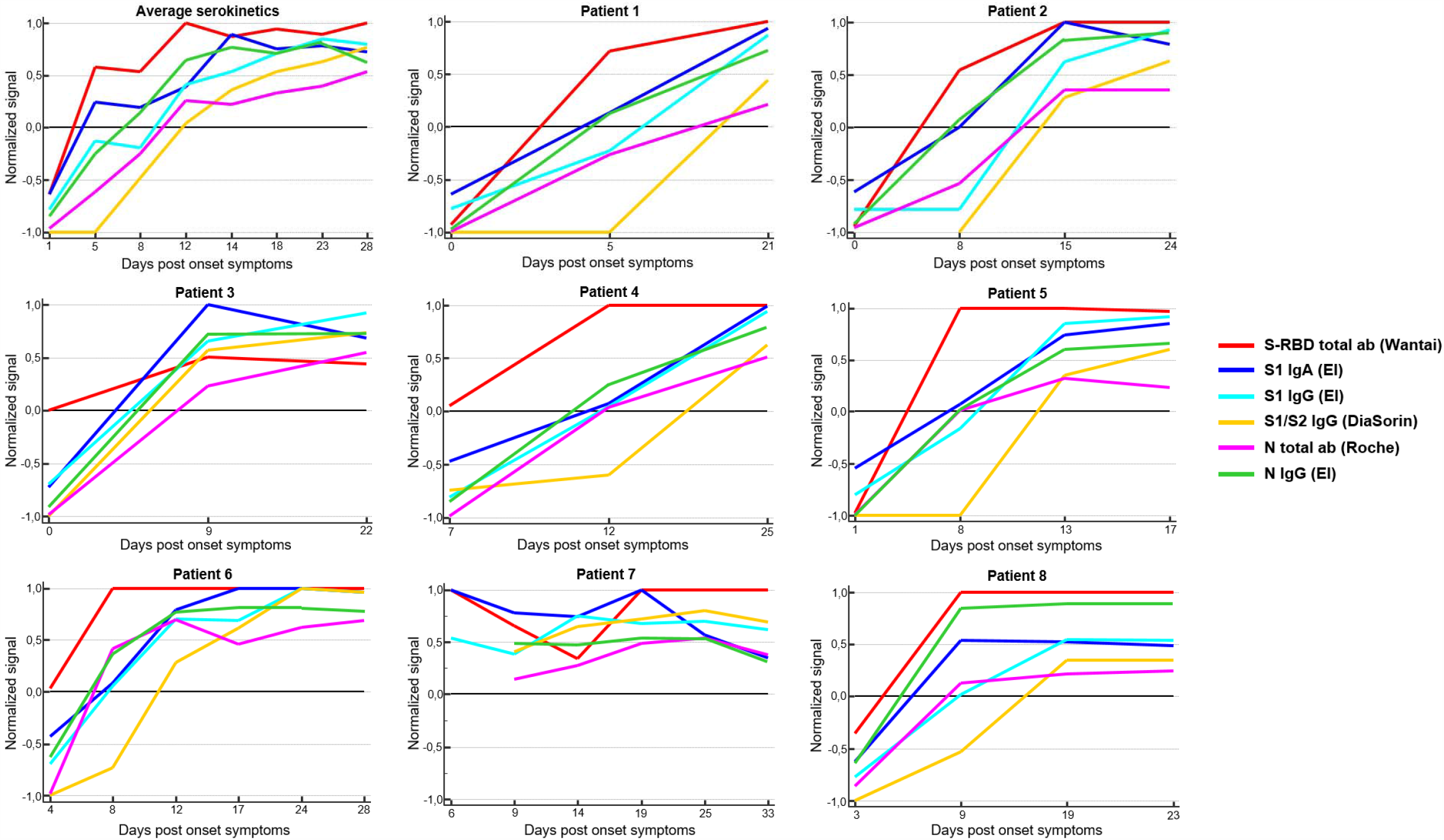
Kinetics of seroconversion in critically ill COVID-19 patients. The upper left panel shows the average kinetics of seroconversion in 13 intensive care unit patients. The other panels show the kinetics in 8 individual patients for whom 3 or more data points were available. Graphs represent for each of the indicated serology tests the normalized signal over time, fitted to a scale from -1 to +1 with 0 (black line) representing the assays’ cutoff as described in Statistical Analysis.

**Figure 2.**
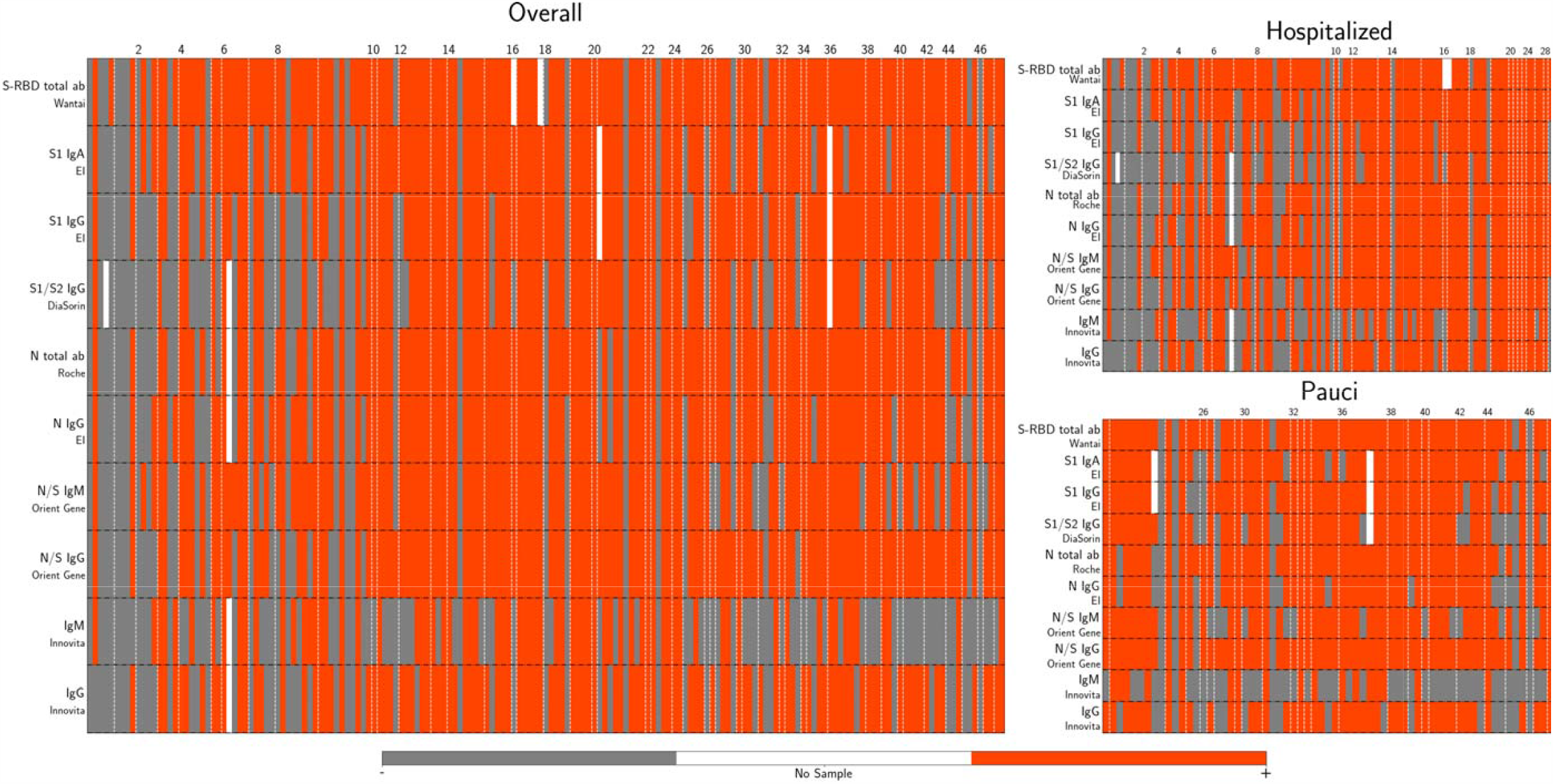
Heatmap of humoral immune response to SARS-CoV-2 in individual samples. Visualization of the concordance of presence or absence of antibodies to RBD/S1/S2/N proteins across the various assays in all samples and grouped according to disease severity as a function of the indicated number of days after symptom onset (top, X-axis). Orange boxes indicate a positive result of the indicated assay, gray boxes are negative results and white boxes were not measured (sample exhausted). From top to bottom: total antibodies to S-RBD (Wantai SARS-COV-2 Ab ELISA), IgA to S1 (EI, EUROIMMUN Anti-SARS-CoV-2 IgA), IgG to S1 (EI, EUROIMMUN Anti-SARS-CoV-2 IgG), IgG to S1/S2 (Diasorin, LIAISON SARS-CoV-2 S1/S2 IgG), total antibodies to N (Roche, Elecsys Anti-SARS-CoV-2 assay), IgG to N (EI, EUROIMMUN Anti-SARS-CoV-2-NCP(IgG)), IgM and IgG to N/S (Orient Gene COVID-19 IgG/IgM Rapid Test), IgM and IgG to undisclosed epitope (Innovita 2019-nCoV Ab rapid test). The heatmap was created in Python 3.7.7.

## Results

### Analytical specificity

The specificity was evaluated on 57 pre-pandemic samples from individuals infected with other HCoV viruses (229E/HKU1/OC43), other infectious agents or with positivity to anti-nuclear factor or rheumatoid factor (detailed in Supplementary Table 1). The Wantai SARS-COV-2 Ab ELISA, Elecsys Anti-SARS-CoV-2 assay, EUROIMMUN Anti-SARS-CoV-2 IgG and Innovita 2019-nCoV Ab Test all achieved 100% specificity (Table 1). The EUROIMMUN Anti-SARS-CoV-2 IgA and Orient Gene COVID-19 IgG/IgM Rapid Test showed the lowest specificity (91.1% and 92.9% respectively) and were the only to cross-react with common cold HCoV viruses. LIAISON SARS-CoV-2 S1/S2 IgG (96.4% specificity) was the only to show interference by rheumatoid factor (Supplementary table 1).

**Table 1.**
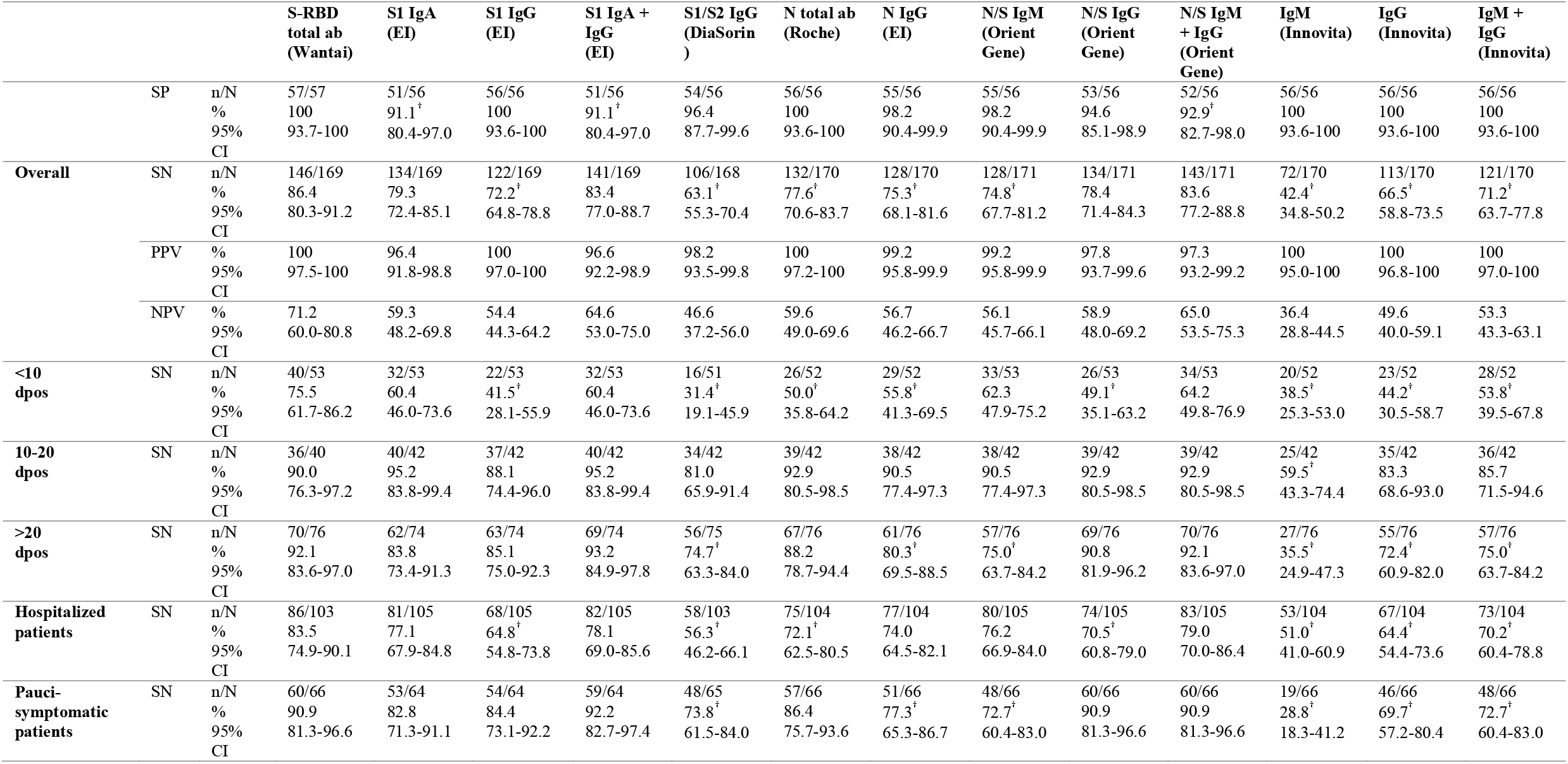
Performance characteristics of serology kits versus the result of PCR. Sensitivities (SN) were expressed as percentage of samples showing detectable antibodies with the indicated serology test, assuming seroconversion in all patients with patients with PCR-confirmed SARS-CoV-2 infection. The table shows sensitivity separately in the hospitalized and paucisymptomatic cohorts and both combined (overall). Data were additionally categorized in three timeframes: less than 10 days post onset of symptoms (dpos), between 10 and 20 dpos and more than 20 dpos. Specificities (SP) were measured on pre-pandemic samples. Medcalc’s (version 12.2.1, Belgium) diagnostic test (2×2) was used for calculation of sensitivity, specificity, positive predictive value (PPV) and negative predictive value (NPV). Proportions for categorical variables were compared using χ2 test. † Indicates differences with the Wantai SARS-COV-2 Ab ELISA for which P values less than .05 were considered statistically significant.

### Analytical sensitivity

#### Study participants

Analytical sensitivities were compared on 171 samples obtained from 135 subjects, all with PCR-confirmed SARS-CoV-2 infections, pooled or grouped in two distinct cohorts: hospitalized and paucisymptomatic COVID-19 patients. *Hospitalized patients* included 105 samples from 71 patients who were hospitalized for severe COVID-19 disease, all with a very high level of suspicion of COVID-19 pneumonia on chest CT (CO-RADS score =5) (11): 48 males (median age 65 years, IQR 53-80) and 23 females (median age 9 years, IQR 67-86). Serum samples ranged from 0 to 39 days after patient-reported symptom onset. *Paucisymptomatic patients*: 66 samples from 64 healthcare workers with mild (n=61) or no (n=3) WHO-listed COVID-19 symptoms: myalgia (present in 62.5%), fever (60.9%), dry cough (56.2%), dyspnea (40.6%), severe fatigue (35.9%), headaches (30.0%), loss of smell or taste (26.6%) or diarrhea (18.8%). None of these patients were hospitalized. Serum samples ranged from 11 to 54 days after patient-reported symptom onset.

#### Analytical sensitivity

was evaluated in two ways, in all samples pooled and separately for the hospitalized and paucisymptomatic patients. First, versus SARS-CoV-2 PCR (100% of samples from PCR+ patients) as reference, by measuring the percentage of samples showing antibody titers above the respective assay’s cutoff (Table 1). Second, by comparing each individual assay to the consensus outcome of the majority of seven tested assays (Table 2). The results were congruent. The Wantai SARS-COV-2 Ab ELISA showed the highest overall sensitivity: 86.4% (95%CI 80.3-91.2) versus PCR and 100% (95%CI 97.3-100) versus the consensus result at all time points in in both patient cohorts. Its sensitivity was significantly higher (P<0.05) than all other assays with exception of the Orient Gene COVID-19 IgG/IgM Rapid Test and the EUROIMMUN Anti-SARS-CoV-2 IgG and IgA combined. In a real-world clinical setting, serology assays will be mostly used at later time stages e.g. more than 20 days after symptom onset or to document past SARS-CoV-2 infection in paucisymptomatic patients: in these patients four assays show clinically acceptable sensitivity above 95% versus the consensus result (Table 2): Wantai SARS-COV-2 Ab ELISA, Elecsys Anti-SARS-CoV-2 assay, EUROIMMUN Anti-SARS-CoV-2 IgG and IgA combined and Orient Gene COVID-19 IgG/IgM Rapid Test. The LIAISON SARS-CoV-2 S1/S2 IgG showed a significantly (P<0.05) lower sensitivity compared to all other assays: with a sensitivity versus consensus (Table 2) of 83.6% (95%CI 72.5-91.5) at > 20 days post symptom onset and of 84.2% (95%CI 72.1-92.5) in paucisymptomatic patients, performance is suboptimal. Also, the EUROIMMUN Anti-SARS-CoV-2-NCP(IgG) and Innovita 2019-nCoV Ab Test suffered from limited sensitivity.

**Table 2.**
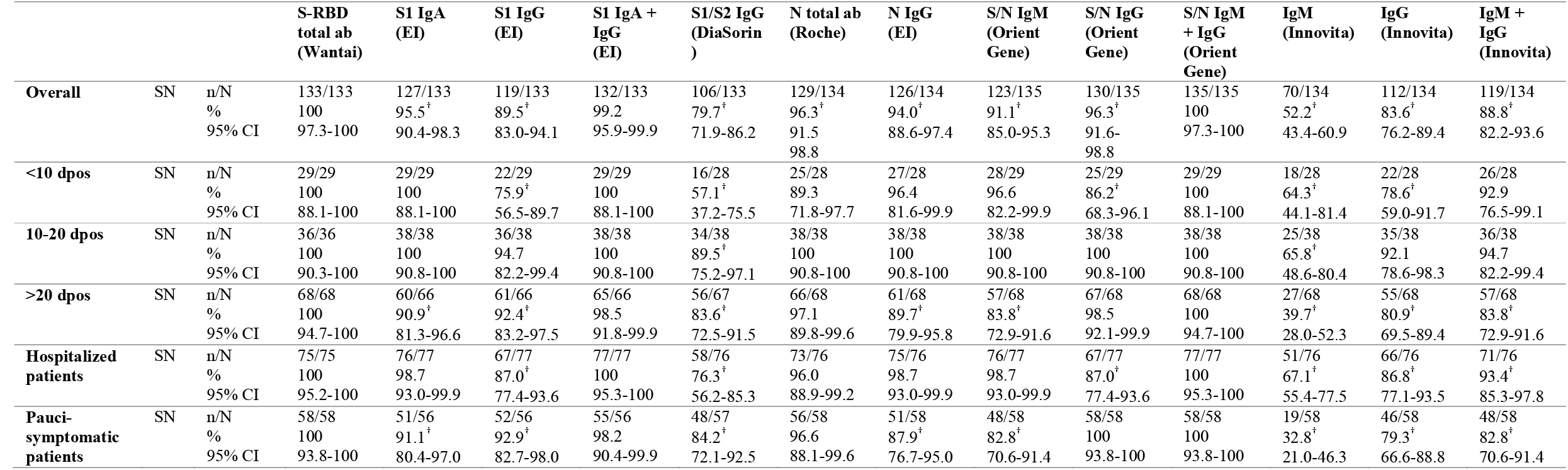
Performance characteristics of serology kits versus the consensus result of all assays. The outcome of the serology tests was compared to the consensus result obtained by the majority of the evaluated assays. Overall sensitivity (SN) was assessed by combining hospitalized and paucisymptomatic cohorts. Data were categorized in three timeframes, in less than 10 days post onset of symptoms (dpos), between 10 and 20 dpos and more than 20 dpos. Medcalc’s (version 12.2.1, Belgium) diagnostic test (2×2) was used for calculation of sensitivity. Proportions for categorical variables were compared using χ2 test. † Indicates differences with the Wantai SARS-COV-2 Ab ELISA for which P values less than .05 were considered statistically significant.

#### Kinetics of seroconversion

We directly compared the kinetics of seroconversion of the ELISA/CLIA assays on consecutive blood samples of 8 patients admitted to intensive care units (Figure 1): in all 8 patients, the RBD-targeting Wantai SARS-COV-2 Ab ELISA was the first to detect seroconversion, followed by the S1-targeting EUROIMMUN Anti-SARS-CoV-2 IgA. Of the N-targeting assays, the EUROIMMUN Anti-SARS-CoV-2-NCP(IgG) detected seroconversion more rapidly than the Elecsys Anti-SARS-CoV-2 assay. The LIAISON SARS-CoV-2 S1/S2 IgG typically was the last to detect seroconversion. The kinetics were additionally studied by a pooled analysis in samples from different patients, grouped according to the timeframe after symptom onset ranging from < 10 days, 10 to 20 days or more than 20 days post onset of symptoms (dpos) (Table 1-2). All tests except Wantai SARS-COV-2 Ab ELISA showed a significantly higher positivity rate between 10 and 20 dpos as compared to less than 10 dpos (P<0.05). No significant differences were observed in positivity rates between 10 and 20 and more than 20 dpos (Table 1) indicating that serology testing can be performed starting from 10 days after symptom onset. In samples less than 10 days after symptom onset, all from hospitalized patients, the Wantai SARS-COV-2 Ab ELISA outperformed all other assays, with a sensitivity of 75.5% (95%CI 61.7-86.2) versus PCR and 100% (95%CI 88.1-100) versus consensus that was however significantly lower than its performance in samples > 20 days dpos (P<0.05).

#### Concordance analysis of humoral immune response on individual samples

We visually plotted the concordance of presence or absence of antibodies to RBD/S1/S2/N proteins across the various assays in all samples and grouped according to disease severity as function of time post symptom onset (Figure 2). For the assays with acceptable overall sensitivity > 95% (Wantai SARS-COV-2 Ab ELISA, Elecsys Anti-SARS-CoV-2 assay, EUROIMMUN Anti-SARS-CoV-2 IgG and IgA combined and Orient Gene COVID-19 IgG/IgM Rapid Test) a good overall concordance was seen in samples > 10 dpos, with 87.7% and 3.5% of samples positive or negative respectively with all four methods. No clear differences were observed in the kinetics of appearance of antibodies to S or N epitopes. Beyond 10 days, only 1.4% (1/71) of hospitalized and 4.7% (3/64) paucisymptomatic patients developed no antibodies.

#### Screening of healthcare workers with suspected SARS-CoV-2 infection

Finally we selected the most performant serology test (Wantai SARS-COV-2 Ab ELISA) to screen a cohort of 84 healthcare workers who failed to obtain a PCR test during peak infection but retrospectively self-reported following COVID-19 symptoms: myalgia (present in 23.8%), fever (21.4%), dry cough (29.8%), dyspnea (20.2%), severe fatigue (14.3%), headaches (5.6%), loss of smell or taste (9.5%) or diarrhea (14.3%): 26.2% showed detectable antibodies as compared to national survey data of 8.4% in unselected healthcare workers and 4.3% in the healthy blood donors at in the same timeframe (Sciensano, Belgian serosurveillance data, sampling from May 6-10, 2020).

## Discussion

In this study we report on the performance characteristics of seven commercially available serology tests for detection of antibodies against the SARS-CoV-2 S protein (S-RBD total antibodies, S1/S2 IgG, S1 IgA and IgG), the N protein (N total antibodies, N IgG), and both proteins (N/S IgM and IgG). This study is the first to report performance of Elecsys Anti-SARS-CoV-2 assay on the Cobas e601 module. We specifically investigated their relative value as a complementary diagnostic tool to screen for prior SARS-CoV-2 infection in individuals that were not (conclusively) tested by PCR in early stage of active viral replication up to 10 days after onset of symptoms. As working definition for acceptable performance, we propose that such assay should combine a minimal sensitivity of 95% versus a consensus estimate and a high specificity above 98% in samples taken 20 days or more after symptom onset, also in subjects who experienced mild SARS-CoV-2 symptoms. Based on these criteria, the Wantai SARS-COV-2 Ab ELISA, the Elecsys Anti-SARS-CoV-2 assay and the Innovita 2019-nCoV Ab Test all showed acceptable specificity. In terms of sensitivity versus the consensus result obtained by all tests, the Wantai SARS-COV-2 Ab ELISA, the Elecsys Anti-SARS-CoV-2 assay the EUROIMMUN Anti-SARS-CoV-2 IgG combined with IgA and the Orient Gene COVID-19 IgG/IgM Rapid Test are acceptable. Overall, only the Wantai SARS-COV-2 Ab ELISA and the Elecsys Anti-SARS-CoV-2 assay reached the proposed acceptance criteria, with the Wantai SARS-COV-2 Ab ELISA clearly outperforming all other evaluated assays.

A strength of our study is that the parallel evaluation of several kits allowed a reliable direct comparison of diagnostic performance using the cutoffs provided by the manufacturers. Also, our patient cohorts, including not only severe COVID-19 patients but also a sizeable cohort of mild SARS-CoV-2 infections provides a good estimate on the assays’ performances in the intended target population. We observed no notable differences in the rate of seroconversion between severe and milder SARS-CoV-2 infections, nor in its timing.

The limitations of our study are that the specificity analysis requires further extension, particularly with time series analyzing false positive seroconversion triggered by other HCoV (229E/HKU1/OC43/NL63) during the common cold season, and that our study did not include a sizeable cohort of fully asymptomatic SARS-CoV-2 infections. Our study also focused on qualitative analysis and did not investigate differences in assays’ performance for quantification of antibody titers. In critically ill COVID-19 patients, SARS-CoV-2 antibody levels were reported to correlate to disease severity (4) by triggering bradykinin and complement activation pathways. The assays evaluated here show large variations in their dynamic range (raw data in Supplementary Information), ranging from a good linearity for the LIAISON SARS-CoV-2 S1/S2 IgG (12) to a limited dynamic range with rapid signal saturation for the most sensitive: with a sample volume input of 100 µL that is 10-20 times higher than the other evaluated assays, the Wantai SARS-COV-2 Ab ELISA is clearly designed towards high sensitivity by maximal antibody capture. Caution is thus warranted when comparing (semi)quantitative estimates of antibody titers across platforms before certified standards with known titers become available.

Our data are compatible with other cross-platform evaluations (13) indicating superior performance of the Wantai SARS-COV-2 Ab ELISA as compared to EUROIMMUN Anti-SARS-CoV-2 IgG and IgA. Our data are, however, discrepant with another study reporting a sensitivity of 100% and 99% specificity for the LIAISON SARS-CoV-2 S1/S2 IgG (12), obtained on a small set of 125 samples including only 40 PCR-confirmed patients and after ROC-optimization of assay cutoffs. Since we observed considerable lot-to-lot variations in the raw signals of the two LIAISON SARS-CoV-2 S1/S2 IgG kits tested, we feel that caution is warranted and cutoffs should only be optimized on better powered data sets and proper assessment of different lots.

It was reported that antibodies against S protein appear later in infection than antibodies against the N protein (4,7). We also observed faster seroconversion of N-versus S1-targeting IgG in the EUROIMMUN assays. On the other hand, we also observed a much faster seroconversion of total antibodies (IgA/IgM/IgG) to S-RBD (Wantai) than N-protein (Elecsys). Within the same epitope/assay format (EUROIMMUN to S1), IgA antibodies clearly precede IgG. IgM does not precede IgG as evident from both rapid tests. Overall, our data suggest that speed of seroconversion depends more on assay design, recombinant viral epitope and antibody isotypes covered, and that overall sensitivity is likely enhanced when both IgA and IgG isotypes are measured.

In conclusion, our study supports the clinical use of both Wantai SARS-COV-2 Ab ELISA and Elecsys Anti-SARS-CoV-2 assay for sensitive and specific screening of SARS-CoV-2 antibodies from 10 days after symptom onset. Within 10 days after symptom onset, only Wantai SARS-COV-2 Ab ELISA achieves medically relevant diagnostic power.

## Data Availability

Source raw data are shared as online supplementary file with the article

## Author statements on competing interests and funding

The authors declare no conflict of interest. This work was supported by a private donation by board members of Fagron (Nazareth, Belgium), a healthcare company, to RADar, the teaching and education initiative of AZ Delta General Hospital, to be used as unconditional research grant for data collection, collaborative collaboration and open access publication. The sponsor had no influence on the study design, data interpretation and drafting of the manuscript.

## Author contribution statement and data sharing statement

Study design: GM, ASDC, DDS, PH. Statistical data analysis: PH, ASDC, DDS. Data interpretation: PH, GM, ASDC, DDS. Data visualization: KH. Data collection: PH, ASDC. Manuscript preparation, lead: PH, GM, ASDC. Manuscript, supportive: KH, DDS. GM is guarantor of the study. Source data discussed in the paper are available on email request to the guarantor of this study.

## Notes

### Competing Interest Statement

The authors have declared no competing interest.

### Clinical Trial

This study was not registered in ClinicalTrials.gov since it is an observational diagnostic accuracy study. All necessary IRB approvals were obtained.

### Author Declarations

The study was approved by the AZ Delta ethical committee with a waiver of informed consent from the hospitalized COVID-19 patients (Clinical Trial Number IRB B1172020000009) and with written informed consent from participants with paucisymptomatic and suspected SARS-CoV-2 infections (Clinical Trial Number B1172020000006).

